# Determinants of Hypertension Prevention Practices Among Adults in a High-Density Residential Area in the Capital-City: A Cross-Sectional Study in Mtendere Compound, Lusaka, Zambia

**DOI:** 10.64898/2026.02.18.26346537

**Authors:** Robert Mwale, Jeff Mugoba, John Soko, Larry L. S. Mooka, Mukumbuta Nawa

**Author notes:** **Corresponding Author** Robert Mwale. Levy Mwanawasa Medical University, School of Public health and Environmental studies. P. O. Box 33991, Lusaka, Zambia.

## Abstract

**Background:** Hypertension is a growing contributor to morbidity and mortality in sub-Saharan Africa, yet many adults are still unaware of how to prevent it. In Zambia, preventive behaviours such as regular exercise, reduced salt intake, and routine blood pressure checks are underutilised. This study examined the factors associated with hypertension prevention practices among adults in Mtendere Compound, a high-density residential area in the capital-city of Zambia Lusaka.

**Methods:** A cross-sectional study was conducted among 382 adults aged 18 to 65 years, selected through multistage random sampling. Data were collected using structured interviewer-administered questionnaires adapted from the WHO-STEPS tools. Key variables included demographic characteristics, knowledge of hypertension, health-seeking behaviour, and lifestyle factors. Descriptive statistics and logistic regression were used to analyse associations between independent variables and hypertension prevention practices as a compound outcome variable including low salt intake, regular exercises and check-ups.

**Results:** Among participants, 58.6% showed adequate knowledge of hypertension prevention. However, only 41.1% reported practising at least three recommended behaviours. Receiving health advice at a facility (AOR: 3.09, 95% CI: 1.72-5.56), having good knowledge (AOR: 2.64, 95% CI: 1.46-4.75), and being employed (AOR: 1.78, 95% CI: 1.02-3.09) were independently associated with practicing preventive behaviours. Age, sex, and education level were not statistically significant predictors.

**Conclusion:** While majority of the respondents had adequate knowledge of hypertension prevention, the actual practice was low. This study underscores the community-based reinforcement of social behavioural change (SBC) to transform knowledge into practice among at risk populations. Specifically, the study recommends tailoring interventions on hypertension prevention to reach unemployed and under-informed populations informal settlements and other high-density settings in major towns and cities in sub-Saharan Africa.

## Introduction

Hypertension, or high blood pressure, is a major global health concern affecting over 1.28 billion people worldwide, with two-thirds of cases occurring in low-and middle-income countries (WHO, 2024). In sub-Saharan Africa, hypertension is a significant health burden, with prevalence rates around 18.6% in urban areas (Doulougou et al., 2014). Despite its asymptomatic nature in initial stages, hypertension is still a leading cause of cardiovascular disease, stroke, and kidney failure (WHO, 2024). In sub-Saharan Africa, the burden of hypertension is growing rapidly due to demographic transitions, urbanisation, poor diets, and declining physical activity (Matemane et al., 2024; Cavagna et al., 2021). Zambia, like many other countries in sub-Saharan Africa, is experiencing a double burden of endemic infectious diseases and rising incidence of chronic Non-Communicable Diseases (Musonda et al., 2024). The urban population, particularly in high-density areas such as Mtendere Compound in Lusaka, is increasingly exposed to lifestyle-related risk factors, including high salt intake, sedentary living, and alcohol consumption (Goma et al., 2019; Ng’ambi et al., 2022). However, existing literature in Zambia tends to focus on the prevalence and treatment of hypertension rather than on preventive behaviours among at-risk urban populations.

Existing studies from rural Zambia report a high prevalence of hypertension, but underdiagnosis is still a significant issue due to poor health-seeking behaviour and limited access to care (Rush et al., 2018; Mahdi et al., 2021). There is a pressing need for evidence that focuses not only on treatment but on prevention. Furthermore, there is a gap in the literature examining the factors influencing preventive practices among adults in peri-urban Zambia, where socioeconomic challenges and lifestyle shifts converge (Mweemba et al., 2011, Nawa et al, 2024). Understanding these determinants is critical for informing targeted public health interventions, especially in informal settlements where hypertension often goes unnoticed and unmanaged. This study, therefore, aimed to identify the factors associated with the practices of hypertension prevention among adults aged 18 to 65 years living in Mtendere compound, Lusaka. By focusing on prevention rather than treatment, it contributes to a shift in how urban hypertension can be addressed not only in Zambia but in similar settings in sub-Saharan Africa where there are high-density and informal settlements in major cities and towns.

## Methods

### Study Design and Setting

This study employed a cross-sectional design. Mtendere compound is characterised by high population density, informal housing, and limited access to basic services such as water and sanitation. It also faces a growing burden of non-communicable diseases, including hypertension, linked to urban lifestyle shifts such as increased consumption of processed foods, reduced physical activity, and high levels of unemployment.

### Study Population and Sampling

The study population included adults aged 18 to 65 years who were permanent residents of Mtendere. A multistage sampling approach was used. First, the compound was divided into five sections; within these, thirty clusters were initially mapped, from which fifteen clusters were randomly selected. Households within each selected cluster were then selected using systematic random sampling to identify eligible participants.

### Sample Size Determination

The required sample size was calculated using the Kelsey two-population formula to detect a difference of at least 15% (Kelsey et al, 1996). Assuming a 32% prevalence of hypertension in the unexposed group and a detectable difference of 15%, with a 95% confidence level and 80% power, the minimum sample size was 258 (Rush et al, 2018). This was adjusted by 20% to account for non-response and multistage sampling resulting in a final sample size of 305 participants.

### Data Collection Tools and Procedures

Data were collected using a structured interviewer-administered questionnaire adapted from the WHO STEPS instrument for non-communicable disease surveillance (Goma et al 2019). The tool captured demographic information, knowledge and attitudes towards hypertension, health-seeking behaviour, and lifestyle practices such as diet, smoking, alcohol use, and physical activity. The questionnaire was pre-tested for clarity and consistency before use and administered in Chinyanja, the dominant local language, by trained research assistants.

### Data Management and Analysis

The collected data were entered into Microsoft Excel, cleaned for accuracy, and then exported into STATA 16 for analysis. Descriptive statistics were used to summarise participant characteristics and prevention behaviours using frequencies and percentages. Bivariate associations between prevention practices and independent variables were evaluated using Chi-square or Fisher’s exact tests, as appropriate. Variables with p-values <0.05 in bivariate analysis were included in multivariate logistic regression to identify independent predictors of hypertension prevention practices.

### Ethical Considerations

Ethical approval was obtained from the University of Lusaka Research Ethics Committee (Approval No. FWA00033228-480/ (08)/ (08)/ (2024). Informed consent was secured from each participant after explanation of the study objectives and procedures. To protect privacy, data were anonymised using coded identifiers, and completed questionnaires were stored securely under lock and key. Participants retained the right to withdraw at any stage without any consequence.

## Results

A total of 305 participants were recruited for this study. The mean age was 38.4 years (SD ± 11.2), with most of the participants (65%) aged between 25 and 49 years. Slightly more than half were male (52%), and over half (51%) were married. The proportion of participants diagnosed with hypertension was 42% (129/305), but only 71% of those reported being on medication at the time of the study. Table 1 shows the basic characterises of the respondents.

**Table 1.** Prevalence of Hypertension and Socio-Demographic Characteristics.

| Variable | Options | Frequency | Percentage |
| --- | --- | --- | --- |
| Sex | Female | 146 | 48 |
|  | Male | 158 | 52 |
| Age (Years) | 18 - 24 | 40 | 13 |
|  | 25 - 49 | 198 | 65 |
|  | 50 - 59 | 46 | 15 |
|  | 60 - 75 | 21 | 7 |
| Marital Status | Single | 103 | 34 |
|  | Married | 151 | 51 |
|  | Separated | 19 | 6 |
|  | Divorced | 14 | 5 |
|  | Widowed | 12 | 4 |
| High Blood Pressure | Yes | 129 | 42 |
|  | No | 176 | 58 |
| High BP and On Treatment | Yes | 92 | 71 |
|  | No | 37 | 29 |

### Lifestyles Characteristics and Habits

Majority 79% (241/305) of the respondents did not smoke tobacco products, only 21 percent smoked cigarettes and other tobacco products and a further 10% used smokeless tobacco products such as vaping. And 90% of the respondent did not use any smokeless products or smoked any tobacco products. On alcohol use behaviour the study showed that 53% did not use alcohol whilst the rest of 47% showed alcohol use behaviour. Furthermore, the study showed that at least 69% (207/305) ate fruits every week. And 31%(95/305) of the respondent said no to eating fruits. The study also showed that 49% (150/305) said they limit eating processed foods compared to 51% (154/305) which showed that they did not limit eating processed foods. On spice use the study reveals that 30% (90/305) responded yes to the use of the spice whilst the rest of the 70% (214/305) said no to the use of spices in their foods. Table 2 shows other characteristics and habits that affect hypertension.

**Table 2:** Lifestyle Characteristics and Habits.

| Variable | Options | Frequency | Percentage |
| --- | --- | --- | --- |
| Smoke Tobacco Products | No | 241 | 79 |
|  | Yes | 64 | 21 |
| Smokeless Tobacco | No | 276 | 90 |
|  | Yes | 29 | 10 |
| Alcohol | No | 162 | 53 |
|  | Yes | 143 | 47 |
| Eat Fruits every week | No | 95 | 31 |
|  | Yes | 207 | 69 |
| Limits Processed Foods | No | 154 | 51 |
|  | Yes | 150 | 49 |
| Use Spices | No | 214 | 70 |
|  | Yes | 90 | 30 |
| Avoid Food Outside home | No | 183 | 60 |
|  | Yes | 121 | 40 |
| High Labour Work | No | 236 | 78 |
|  | Yes | 67 | 22 |
| Moderate Labour Work | No | 257 | 85 |
|  | Yes | 46 | 15 |
| Walk to Places | No | 131 | 43 |
|  | Yes | 172 | 57 |
| Exercises | No | 250 | 83 |
|  | Yes | 53 | 17 |
| History of Stroke | No | 288 | 94 |
|  | Yes | 17 | 6 |
| Take Aspirin to Prevent Stroke | No | 272 | 90 |
|  | Yes | 31 | 10 |
| Take Cholesterol Lowering drugs | No | 290 | 95 |
|  | Yes | 15 | 5 |

### Hypertension Prevention Knowledge and Practices

A total of 58.6% (179/305) of participants reported knowing at least three prevention strategies, only 41% consistently practiced them. Fruit consumption and physical activity were the most inconsistently followed behaviours. Among those with adequate knowledge, preventive practice was significantly more common (*p < 0*.*001*). Table 3 shows the summary of some hypertension prevention practices.

**Table 3.**
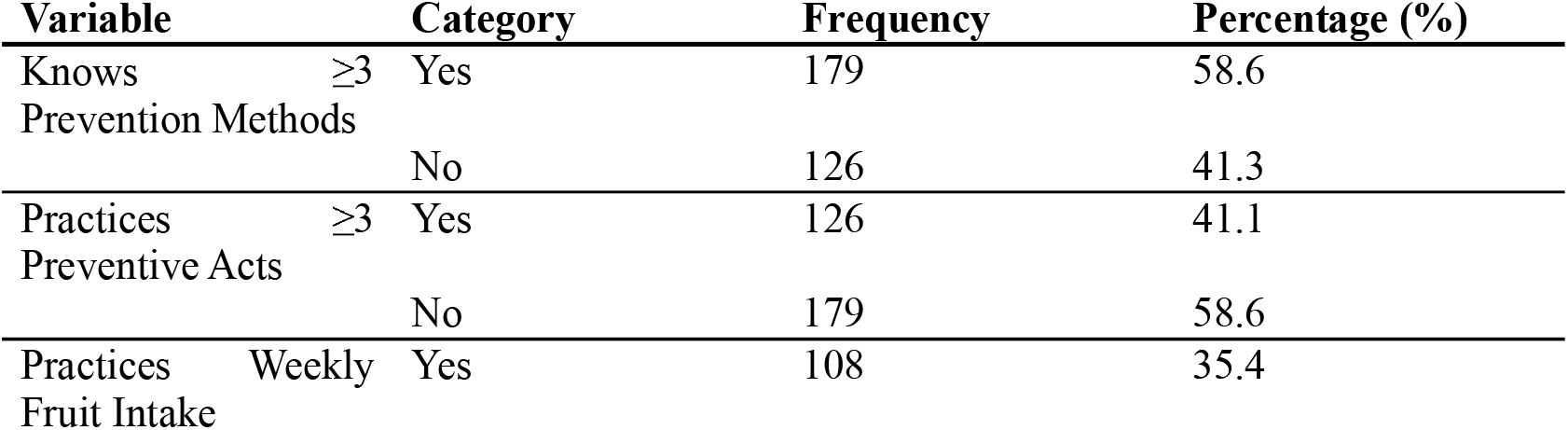
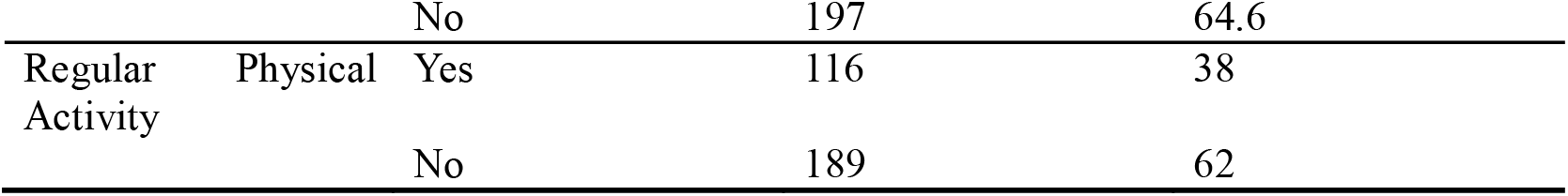
Knowledge and Practice of Hypertension Prevention (n = 305)

**Table 4.** Sociodemographic Characteristics and Preventive Practices Stratified by Hypertension.

| Variable | Options | No High BP n(%) | High BP n(%) | P-Value |
| --- | --- | --- | --- | --- |
| Sex | Female | 75 (51) | 71 (49) | 0.036* |
|  | Male | 100 (63) | 58 (37) |  |
| Age (Years) | 18 – 24 | 33 (83) | 7 (17) | < 0.001** |
|  | 25 – 49 | 124 (63) | 74 (37) |  |
|  | 50 – 59 | 17 (37) | 29 (63) |  |
|  | 60 – 75 | 2 (10) | 19 (90) |  |
| Marital Status | Single | 79 (77) | 24 (23) | < 0.001** |
|  | Married | 69 (46) | 82 (54) |  |
|  | Separated | 14 (74) | 5 (26) |  |
|  | Divorced | 8 (57) | 6 (42) |  |
|  | Widowed | 3 (25) | 9 (75) |  |
| Employment Status | Formal Employed | 60 (53) | 53 (47) | < 0.001** |
|  | Retired | 1 (8) | 11 (92) |  |
|  | Self-Employed | 37 (58) | 27 (42) |  |
|  | Student | 39 (87) | 6 (13) |  |
|  | Unemployed | 38 (54) | 32 (46) |  |
| Smoke Tobacco Products | No | 151 (63) | 90 (37) | 0.001* |
|  | Yes | 25 (39) | 39 (61) |  |
| Smokeless Tobacco | No | 163 (59) | 113 (41) | 0.140 |
|  | Yes | 13 (45) | 16 (55) |  |
| Alcohol | No | 110 (68) | 52 (32) | < 0.001* |
|  | Yes | 66 (46) | 77 (54) |  |
| Eat Fruits every week | No | 48 (51) | 47 (49) | 0.077* |
|  | Yes | 127 (61) | 80 (39) |  |
| Limits Processed Foods | No | 91 (59) | 63 (41) | 0.669* |
|  | Yes | 85 (57) | 65 (43) |  |
| Use Spices | No | 128 (60) | 86 (40) | 0.296* |
|  | Yes | 48 (53) | 42 (47) |  |
| Avoid Food Outside home | No | 103 (56) | 80 (44) | 0.578* |
|  | Yes | 72 (60) | 49 (40) |  |
| High Labour Work | No | 143 (61) | 93 (39) | 0.036* |
|  | Yes | 31 (46) | 36 (54) |  |
| Moderate Labour Work | No | 151 (59) | 106 (41) | 0.405* |
|  | Yes | 24 (52) | 22 (49) |  |
| Walk to Places | No | 86 (66) | 45 (34) | 0.012* |
|  | Yes | 88 (51) | 84 (49) |  |
| Exercises | No | 144 (58) | 106 (42) | 0.905* |
|  | Yes | 31 (58) | 22 (42) |  |
| History of Stroke | No | 172 (60) | 116 (40) | 0.005** |
|  | Yes | 4 (24) | 13 (76) |  |
| Take Aspirin to Prevent Stroke | No | 173 (64) | 99 (36) | < 0.001** |
|  | Yes | 2 (6) | 29 (94) |  |
| Take Cholesterol Lowering drugs | No | 175 (60) | 115 (40) | < 0.001** |
|  | Yes | 1 (7) | 14 (93) |  |

\*P-value determined using Chi-Square
\*\*P value determined using Fisher's Exact Test

### Sociodemographic Characteristics and Prevention Practices Stratified by Hypertension

Statistical tests revealed significant associations between practicing preventive measures and sex, employment status, smoking, alcohol use, and hypertension knowledge. For instance, non-smokers and those who did not take alcohol were significantly more likely to engage in preventive behaviours (*p < 0*.*001*). Table 3 summarises the potential associations between independent variables and hypertension prevention practices.

### Factors Associated with Hypertension

After controlling for confounders, three variables remained statistically significant in predicting preventable practices among the respondents. Those who were employed (AOR: 1.78; P-value = 0.045), whilst those who had adequate knowledge were also more likely to practice hypertension prevention (AOR: 2.64; P-value <0.001). Further, those who had received advice at health facility were also more likely to practice hypertension prevention (AOR: 3.09; 95% CI: 1.72-5.56). Table 5 shows the factors associated with hypertension prevention.

**Table 5:** Factors Associated with Hypertension Prevention.

| Variable | Options | uOR | P-Value | aOR | P-Value |
| --- | --- | --- | --- | --- | --- |
| Sex | Female | Ref |  | Ref |  |
|  | Male | 0.86 | 0.567 | 1.28 | 0.464 |
| Age (Years) | 18 - 24 | Ref |  | Ref |  |
|  | 25 - 65 | 1.03 | 0.933 | 1.34 | 0.600 |
| Marital Status | Single | Ref |  | Ref |  |
|  | Married | 0.94 | 0.926 | 0.55 | 0.446 |
|  | Separated | 0.47 | 0.346 | 0.38 | 0.309 |
|  | Divorced | 0.44 | 0.239 | 0.32 | 0.180 |
|  | Widowed | 0.82 | 0.83 | 0.68 | 0.738 |
| Employed | No | Ref |  | Ref |  |
|  | Yes | 6.26 | 0.084 | **1.78** | **0.042** |
| Smoke Tobacco Products | No | Ref |  | Ref |  |
|  | Yes | 2.26 | 0.023 | 0.62 | 0.434 |
| Smokeless Tobacco | No | Ref |  | Ref |  |
|  | Yes | 0.957 | 0.312 | 0.96 | 0.957 |
| Alcohol | No | Ref |  | Ref |  |
|  | Yes | 0.99 | 0.261 | 0.76 | 0.351 |
| Eat Fruits every week | No | Ref |  | Ref |  |
|  | Yes | 0.29 | 0.711 | 0.43 | 0.235 |
| Use Spices | No | Ref |  | Ref |  |
|  | Yes | 1.62 | 0.716 | 1.65 | 0.633 |
| Avoid Food Outside home | No | Ref |  | Ref |  |
|  | Yes | 0.86 | 0.401 | 0.91 | 0.374 |
| Adequate Knowledge | No | Ref |  | Ref |  |
|  | Yes | **3.01** | **< 0.001** | **2.64** | **< 0.001** |
| Facility Advice | No | Ref |  | Ref |  |
|  | Yes | **4.32** | **< 0.001** | **3.09** | **< 0.001** |

## Discussion

Overall, the analysis revealed that while basic awareness of hypertension prevention was high among adults in this high density area, this knowledge did not always translate into consistent behaviour, however, adequate knowledge was still significantly associated with hypertension prevention practices. Preventive practices were significantly associated with being in employment, adequate knowledge, and when someone received advice from a health facility. Factors such as age, sex, and educational level, traditionally assumed to be predictors, did not show significant associations, suggesting that behavioural and structural enablers may be more influential than demographic factors alone.

These findings align with the Health Belief Model, which suggests that perceived susceptibility, perceived benefits, and cues to action influence preventive behaviours (Alamer et al, 2024). While participants acknowledged the seriousness of hypertension, the low adoption of consistent preventive behaviours may reflect low perceived vulnerability or low confidence in sustaining lifestyle changes. Notably, the strong association between receiving health advice at a facility and engaging in preventive practices reinforces the importance of provider-driven “cues to action.” However, the low proportion of participants who reported receiving such advice suggests a missed opportunity within routine care to promote NCD prevention. The association between knowledge and behaviour is consistent with findings in Zambia and elsewhere in sub-Saharan Africa, where health literacy has been positively linked to non-communicable disease prevention (Musonda et al., 2024; Mweemba et al., 2011).

Employment status also played a significant role in shaping behaviour. Employed participants were more likely to engage in preventive action due to greater access to information, health services, and nutritious food options. This underscores the importance of addressing social determinants of health, particularly economic empowerment and access to a supportive environment, as part of NCD prevention strategies. In contrast, unemployment may limit access to health services, increase stress levels, and encourage risk behaviours such as smoking or alcohol use, an observation echoed in other studies from urban Zambia (Mahdi et al., 2021).

Interestingly, age, sex, and educational attainment were not significant predictors of behaviours in this study. This differs from findings in rural settings, where older adults and those with lower education levels often exhibit poorer preventive practices. In urban settings like Mtendere, risk exposures, such as poor diet, sedentary lifestyle, and stress, are more uniformly distributed across demographic lines. This may explain why the individual-level differences were less pronounced, and it suggests that urban health strategies must be universally targeted, rather than focused only on high-risk groups. These findings mirror observations from Nulu et. al. (2016), who reported a rising burden of hypertension in urban Zambia irrespective of age group.

The strong predictive value of facility-based health advice highlights a critical and underutilised opportunity. Outpatient visits, which often focus on acute conditions, could be framed to include brief NCD counselling interventions. Embedding these into everyday clinical workforces may increase the reach and impact of hypertension prevention strategies, especially as Zambia expands its universal coverage agenda.

### Limitations

Our study had several limitations. First, it relied on self-reported behaviour, which is subject to social desirability bias, and the cross-sectional design limits causal interpretation. Lastly, the study was conducted in one urban compound, and results may not be generalised to rural areas or other socio-economic settings.

### Implications for Practice and Policy

Urban health systems must integrate prevention counselling into clinical care, particularly at the primary level. Targeted community interventions, such as local health talks, workplace wellness programs, and support for affordable healthy food options, could support individuals in converting knowledge into practice. Importantly, this study adds to a growing body of evidence suggesting that urban NCD risk is socially patterned but broadly distributed, and that effective prevention requires multi-sectoral action beyond the health sector alone.

### Conclusion

This study underscores a persistent gap between knowledge and practice in hypertension prevention among adults in Mtendere, Lusaka. While most participants were aware of key prevention strategies, fewer than half consistently applied them. Employment status, adequate knowledge, and receiving advice from health facilities were significant predictors of preventive behaviour. These findings reflect the influence of both individual beliefs and broader social determinants on health choices. Interventions that focus solely on awareness may fall short unless paired with supportive systems and environments. Embedding NCD prevention messaging into routine clinical care and expanding access to community-based health promotion may help translate knowledge into sustained behaviour change. As Zambia responds to the rising burden of non-communicable diseases, prevention strategies must be holistic, inclusive, and responsive to the realities of urban life.

## Data Availability

All data produced in the present study are available upon reasonable request to the authors

## Declarations

### Consent for Publication

Not applicable. This study does not include any identifiable personal data.

### Availability of Data and Materials

The datasets used and/or analysed during the current study are available from the corresponding author upon reasonable request.

### Authors’ contributions

RM: Conceptualisation, data collection, analysis, original draft writing. JM, JS, LLSM, MN: Manuscript editing, analysis, interpretation of findings, final draft writing and editing. All authors read and approved the final manuscript.

## Acknowledgements

The authors express gratitude to the study participants and the Levy Mwanawasa Medical University for Institutional Support.

